# The Role of Biomarkers in Early Detection of Chronic Disease Risk and Smoking Cessation Efforts among Students, Indonesia

**DOI:** 10.64898/2026.02.19.26346603

**Authors:** Musparlin Halid, Beny Binarto Budi Susilo, Pauzan

## Abstract

**Objective:** The study aimed to analyze factors associated with cotinine levels as an early risk indicator for chronic diseases and students’ readiness to quit smoking in Praya Barat District.

**Methods:** This study used a cross-sectional design involving 563 high school students in Praya Barat District. Data analysis was performed using the Chi-square test and multiple logistic regression to identify determinants of high cotinine levels.

**Results:** A total of 67% of subjects had high cotinine levels, indicating high levels of nicotine exposure among students. The results of the analysis showed that the main determinants of high cotinine levels were cigarette consumption of ≥5 cigarettes/day (AOR=2.426; 95% CI=1.534–3.838; p<0.001), male gender (AOR=2.100; 95% CI=1.358–3.250; p=0.001), family members who smoke (AOR=2.149; 95% CI=1.359–3.399; p=0.001), rarely of exercise (AOR=2.155; 95% CI=1.350–3.440; p=0.001), and personal history of chronic disease (AOR=2.646; 95% CI=1.653–4.234; p<0.001). Meanwhile, willingness to participate in a smoking cessation program did not show a significant relationship (p=0.093).

**Conclusions:** Most students showed high cotinine levels, indicating significant exposure to nicotine and a potential risk of chronic disease in the future. The most influential factors were active smoking behavior, a family environment of smokers, and low levels of physical activity.

## Introduction

Smoking is one of the main risk factors for various chronic diseases, including Chronic Obstructive Pulmonary Disease (COPD), lung cancer, cardiovascular disease, and other chronic diseases (Tao *et al*., 2024). That is a major challenge in efforts to prevent and mitigate the negative impact of tobacco on public health, especially among adolescents (Mahabee-Gittens *et al*., 2021). The habit of smoking that starts at an early age can increase the risk of nicotine addiction and increase the likelihood of various health complications in the future (Singh *et al*., 2021). Based on data from the Central Statistics Agency (CSA) of Indonesia in 2024, it was reported that the prevalence of beginner smokers, especially among teenagers, was 28.99%. Meanwhile, in West Nusa Tenggara (WNT), Indonesia, the prevalence of teenagers who smoke was reported to have reached 32.40%, an increase from the previous year’s figure of only 22.50% (BPS, 2024).

One method that can be used to detect exposure to cigarette smoke and nicotine consumption is biomarker-based intervention (Popescu *et al*., 2023). Biomarkers, such as cotinine, are biological indicators that can be measured in blood or urine to assess an individual’s level of exposure to cigarette smoke, both active and passive (Siddiqi *et al*., 2024). The use of biomarkers not only helps in the early detection of chronic disease risk, but can also be an effective tool to motivate individuals to quit smoking (Land *et al*., 2023).

In the school environment, smoking among students is a serious problem that requires a special approach (McGinnis *et al*., 2022). Several factors that influence smoking behavior in adolescents include peer influence, family environment, and the relative ease of access to tobacco products (Halid *et al*., 2025). Therefore, an appropriate strategy is needed to reduce the number of new smokers, one of which is through the implementation of biomarker-based intervention programs as a form of early detection and education (Fang *et al*., 2023).

Biomarker-based intervention programs in schools have several benefits, including serving as an educational tool for students about the dangers of smoking and providing concrete evidence of the effects of smoking on the body (Sharma *et al*., 2022), and raise individual awareness to quit smoking (Land *et al*., 2022). With biological evidence in the form of cotinine levels or other biomarkers detected in the body (Huque *et al*., 2023), students can be more motivated to reduce or even quit smoking (Ben Fredj *et al*., 2022).

Previous studies have shown that biomarker-based interventions, such as cotinine testing, can improve the effectiveness of smoking cessation programs (Bickel *et al*., 2023). Biomarker-based interventions not only provide objective information about nicotine exposure (Pollock *et al*., 2023), but can also be used as a basis for designing intervention strategies that are more personalized and tailored to each individual’s circumstances (Upadhyay *et al*., 2023). With a more targeted approach, the number of smokers among teenagers can be significantly reduced.

In Praya Barat District, efforts to prevent smoking among high school students still face various challenges, such as a lack of awareness of the long-term effects of smoking and a lack of effective intervention programs at the school level. Therefore, research on biomarker-based interventions (cotinine levels in urine) is needed as a method of early detection of disease risk and as an effort to encourage long-term smoking cessation among students.

The study aims to assess the extent to which biomarker-based interventions (urine cotinine levels) can help identify students exposed to nicotine, both as active and passive smokers. In addition, the study will also evaluate the impact of biomarker-based interventions based on urine analysis results (cotinine levels) in early detection of future chronic disease risks and student motivation to quit smoking. Thus, the results of this study are expected to form the basis for the development of more effective and sustainable policies to be implemented in combating smoking habits among adolescents, particularly in Central Lombok Regency.

## Methods

### Study Design

The study used a cross-sectional design by measuring exposure (cotinine levels, family smoking status, smoking habits) and health outcomes/characteristics at a single point in time (snapshot) among students. The study was conducted at senior high schools in Praya Barat District, Central Lombok Regency, WNT Province, Indonesia, from March to October 2025.

### Participants and Sampling

The study population consisted of all high school students in Praya Barat District, totaling 1,189 students. The sampling technique was performed using Sample Size for Frequency in a Population using the OpenEpi Version 3 calculator at: https://www.openepi.com/SampleSize/SSCohort.htm totaling 563 students who met the inclusion and exclusion criteria. Inclusion criteria included students aged 15-18 years who smoked and were exposed to secondhand smoke and were willing to participate in the study with parental/guardian consent. Exclusion criteria included having a previously diagnosed chronic disease and not completing the study.

### Variables

Cotinine levels (ordinal scale) are defined as the levels of the main metabolite of nicotine in urine, which is formed in the body as a result of exposure to cigarette smoke, either actively or passively, as determined by laboratory testing. Cotinine has a longer half-life than nicotine (16–20 hours), so it is used as a more stable biological biomarker for detecting tobacco exposure in individuals. Cotinine levels in ng/mL (nanograms per milliliter) are categorized as “High” if ≥10 ng/mL, indicating active exposure to nicotine/cigarettes, and “Low” if <10 ng/mL, indicating no or passive exposure to nicotine/cigarettes. Age (ratio scale) is defined as the length of time the respondent has lived since birth until the time of the study, expressed in full years. Age data was obtained through questionnaire completion. Age is presented in the form of Mean and Standard Deviation (SD) to describe the characteristics of the respondents descriptively. Gender (nominal scale) describes the biological identity of respondents, categorized as male and female according to the respondents’ self-identification.

Family member smoking status (nominal scale) is a condition in which one or more family members actively smoke at home. It is categorized as “Yes” if there are family members who smoke and “None” if there are no family members who smoke. Cigarette consumption (ordinal scale) indicates the average number of cigarettes smoked by respondents per day in the past week. Data is categorized based on the intensity of daily cigarette consumption. Categorized as “≥5 cigarettes/day” indicates high consumption and “<5 cigarettes/day” indicates low consumption.

Physical activity (ordinal scale) describes the frequency with which respondents engage in physical activity or exercise in the past week. It is categorized as “Rarely” if the respondent exercises ≥3 times per week for at least 30 minutes per session and “Often” if the respondent exercises <3 times per week or irregularly. History of chronic cough (nominal scale) is defined as the presence of cough complaints lasting ≥3 weeks continuously in the past few months, with or without phlegm. Data was obtained from direct questions to respondents. It is categorized as “Ever” if the respondent has experienced chronic cough and “Never” if the respondent has never experienced chronic cough.

History of shortness of breath (nominal scale) is defined as complaints of breathing difficulties experienced by respondents, especially during activities or at certain times, and have been diagnosed or felt repeatedly. Categorized as “Ever” and “Never”. History of chronic disease (nominal scale) is a condition in which the respondent has been diagnosed by a health professional with a non-communicable disease such as asthma, hypertension, diabetes mellitus, or heart disease. It is categorized as “Ever” and “Never”.

Family history of chronic disease (nominal scale) is defined as the presence of a first-degree relative (father, mother, or sibling) who has been diagnosed with a chronic disease such as diabetes, hypertension, asthma, or heart disease. Categorized as “Yes” indicates that there is a family member with a chronic disease and “None” indicates that there are no family members with chronic diseases. Interest and readiness to quit smoking (ordinal scale) describes the extent to which respondents want to quit smoking in the near future. Categorized as “Yes, very much” indicates that the respondent has a strong desire and high motivation to quit smoking, “Unsure” indicates that the respondent is not yet sure about quitting smoking or is still considering it, and “No” indicates that the respondent has no desire to quit smoking.

Willingness to participate in a smoking cessation program (ordinal scale) describes the respondent’s readiness to participate in smoking cessation intervention activities, such as counseling, mentoring, or training. Categorized as “Yes” indicates that the respondent is willing to participate in a smoking cessation program, “Unsure” indicates that the respondent has not yet decided, and “No” indicates that the respondent is not willing to participate in the program.

### Data Collection

Data collection was conducted using a structured questionnaire containing data on biomarker test results, subject sociodemographics, smoking behavior, chronic disease history, and motivation to quit smoking. Biomarker (urine) health measurements were conducted using a spirometry device to determine the level of exposure to cigarette smoke by testing carbon monoxide (CO) levels in breath, and a urine analyzer to assess nicotine levels in the body. According to Pallant (2020), instruments with a Cronbach’s Alpha value above 70% are considered reliable and suitable for measuring research variables (Pallant, 2020). The results of the questionnaire reliability test revealed that Cronbach’s Alpha value was 0.716 or 72%.

### Data Analysis

Statistical tests were performed using SPSS Version 26. Descriptive analysis was conducted to examine the frequency of the measured variables. Chi-square tests and logistic regression were used to analyze the predictors of chronic disease risk detection and smoking cessation attempts with a p-value <0.05 and Adjusted Odds Ratio (AOR) with 95% Confidence Interval (CI).

### Ethical Approval

The study has been approved by the Health Research Ethics and Academic Integrity Committee (HRE-AIC), University of Bima International MFH with Number: 205/KEPK-IA/VI/2025. Before data collection, subjects were provided with written information regarding the purpose and objectives of the study, and researchers requested consent by signing an informed consent form. Researchers guarantee that subject data will be kept confidential and used solely for research purposes.

## Results

Based on the results of cotinine biomarker testing among students in Praya Barat District, it was found that of the total 563 respondents, 67% had high cotinine levels, while 33% had low cotinine levels (Table 1). The study findings indicate that most students have significant exposure to nicotine, suggesting active smoking habits or passive exposure to cigarette smoke from their surroundings. The proportion of high cotinine levels, which reached more than half of the population, indicates that the level of exposure to tobacco among students is still high, either due to their own smoking behavior or exposure from family members or peers.

**Table 1.** Frequency distribution of cotinine levels (n=563).

| Variables | n (%) |
| --- | --- |
| Cotinine Level | 377 (67) |
| High | 186 (33) |
| Low |  |
| Gender |  |
| Male | 352 (62.5) |
| Female | 211 (37.5) |
| Family member smoking status |  |
| Yes | 386 (68.6) |
| None | 177 (31.4) |
| Cigarette consumption |  |
| $\geq 5$ cigarettes/day | 403 (71.6) |
| $< 5$ cigarettes/day | 160 (28.4) |
| Physical activity |  |
| Rarely | 411 (73) |
| Often | 152 (27) |
| History of chronic cough |  |
| Ever | 394 (70) |
| Never | 169 (30) |
| History of shortness of breath |  |
| Ever | 400 (71) |
| Never | 163 (29) |
| History of chronic disease |  |
| Ever | 401 (71.2) |
| Never | 162 (28.8) |
| Family history of chronic disease |  |
| Yes | 392 (69.6) |
| None | 171 (30.4) |
| Interest and readiness to quit smoking |  |
| Yes, very much | 302 (53.6) |
| Unsure | 190 (33.7) |
| No | 71 (12.6) |
| Willingness to participate in a smoking cessation program |  |
| Yes | 303 (53.8) |
| Unsure | 199 (35.3) |
| No | 61 (10.8) |

In terms of demographic characteristics, the majority of respondents were male (62.5%), while females accounted for 37.5%. Exposure to cigarette smoke in the family environment was also quite high, with 68.6% of respondents having family members who smoked. In terms of smoking behavior, most respondents consumed ≥5 cigarettes per day (71.6%), while 28.4% consumed less than 5 cigarettes per day (Table 1).

Physical activity showed a poor pattern, as 73% of respondents rarely engaged in physical activity and only 27% did so frequently. A history of respiratory disorders was also quite common, with 70% of respondents having experienced chronic cough and 71% having experienced shortness of breath. In addition, 71.2% of respondents had a history of chronic disease, and 69.6% had a history of chronic disease in their family (Table 1).

Regarding behavioral motivation, more than half of the respondents (53.6%) were highly interested and ready to quit smoking, while 33.7% were still undecided and 12.6% had no desire to quit. Willingness to participate in a smoking cessation program showed a similar pattern, with 53.8% stating they were willing, 35.3% feeling hesitant, and 10.8% refusing to participate in the program. These results illustrate that although most respondents have a fairly heavy and risky smoking habit, the level of motivation to quit smoking and participate in cessation programs is still relatively high and could be an opportunity for effective intervention (Table 1).

The results of the bivariate analysis in Table 2 show a number of factors that are significantly associated with cotinine levels in respondents. The average age did not show a significant difference between the high and low cotinine level groups (p=0.801), so age did not affect the difference in cotinine levels. However, gender had a highly significant association (p<0.001), with a much higher proportion of high cotinine levels among males (72.4%) than females (27.6%), while females were more likely to be in the low cotinine level group (57.5%).

**Table 2.**
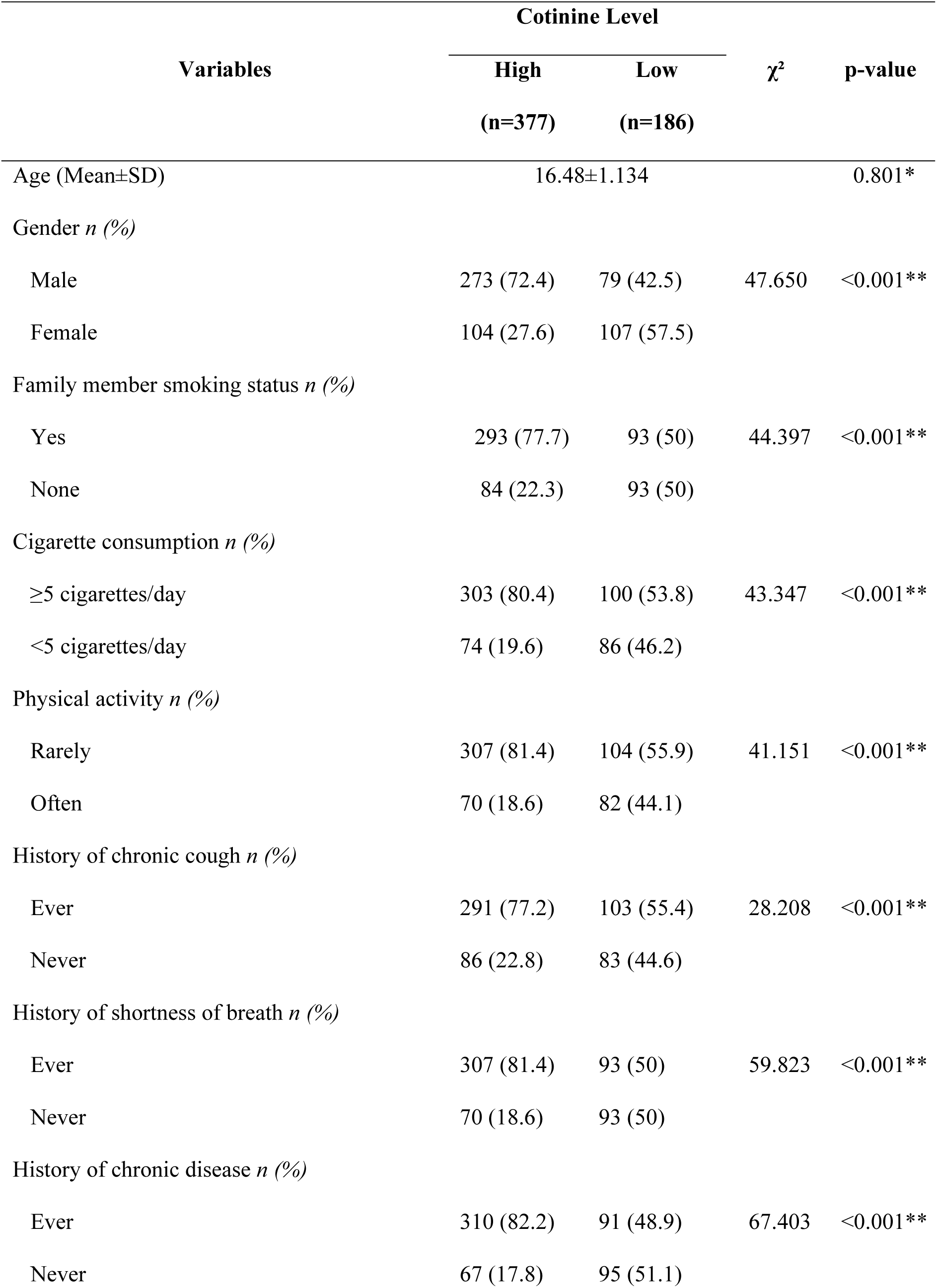
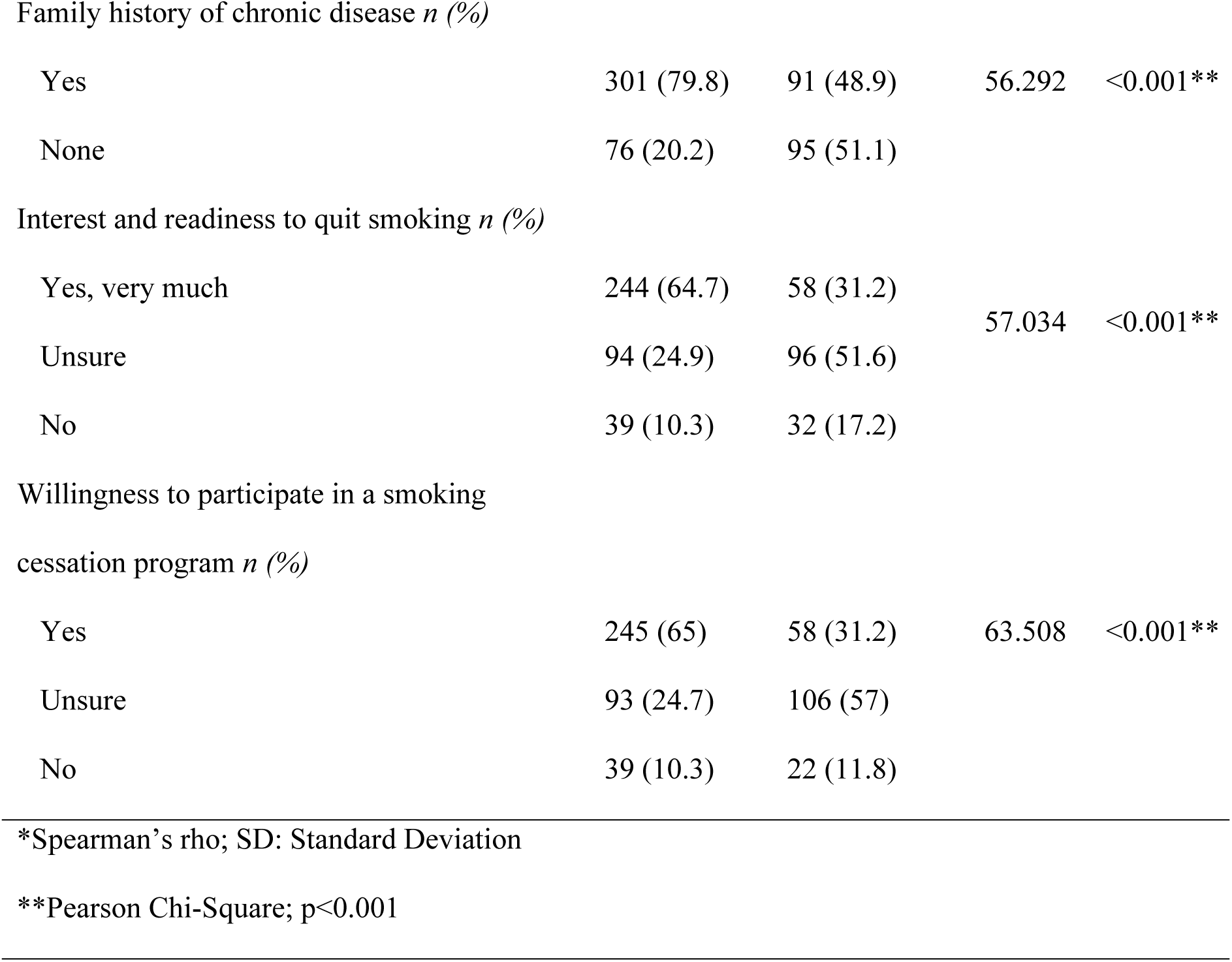
Bivariate analysis of early detection of chronic disease risk and smoking cessation efforts (n=563).

The status of family members who smoke was also significantly associated with cotinine levels (p<0.001). A total of 77.7% of respondents with high cotinine levels had family members who smoked, while the low cotinine level group showed a balanced proportion between those who had and did not have family members who smoked (50% each). Daily cigarette consumption showed a strong association (p<0.001), with 80.4% of respondents with high cotinine levels consuming ≥5 cigarettes per day, while the low cotinine level group consumed <5 cigarettes more frequently (Table 2).

Physical activity also showed a significant association (p<0.001); 81.4% of respondents with high cotinine levels rarely engaged in physical activity, while the low cotinine level group showed a better proportion of physical activity (44.1% engaged in physical activity frequently). A history of respiratory disorders also showed a significant association pattern. Respondents who had experienced chronic cough (77.2%) and shortness of breath (81.4%) were more likely to be in the high cotinine level group than those without such a history (Table 2).

A history of personal and family chronic diseases was also significantly associated with cotinine levels (both p<0.001). A total of 82.2% of respondents with high cotinine levels had a history of chronic disease, and 79.8% had a family history of chronic disease. Conversely, the proportion of respondents without a history of personal or family chronic disease was greater in the low cotinine level group (Table 2).

In terms of behavioral motivation (Table 2), there was a significant association with cotinine levels. Respondents who were very interested in quitting smoking were more likely to be in the high cotinine level group (64.7%), while the low cotinine level group was dominated by respondents who were still undecided (51.6%). A similar pattern was seen in the willingness to participate in a smoking cessation program (p<0.001), with 65% of respondents with high cotinine levels expressing willingness, while the low cotinine level group showed more hesitation (57%).

Overall, various factors related to smoking behavior, physical activity, health history, exposure to family cigarette smoke, and motivation to quit smoking showed a significant association with cotinine levels, indicating that high cotinine levels are closely related to active and passive cigarette exposure and poor health conditions.

The results of the logistic regression analysis in Table 3 show that males are 2.1 times more likely (AOR=2.100; 95% CI: 1.358–3.250; p=0.001) to have high cotinine levels than female students. This indicates that gender is a strong factor influencing nicotine exposure, with males being more vulnerable to engaging in smoking behavior or being in social environments that support this habit. Students who have family members who smoke are 2.15 times more likely to have high cotinine levels (AOR=2.149; 95% CI: 1.359–3.399; p=0.001) than those who do not have family members who smoke. This confirms that exposure to secondhand smoke at home plays an important role in increasing nicotine levels in adolescents.

**Table 3.**
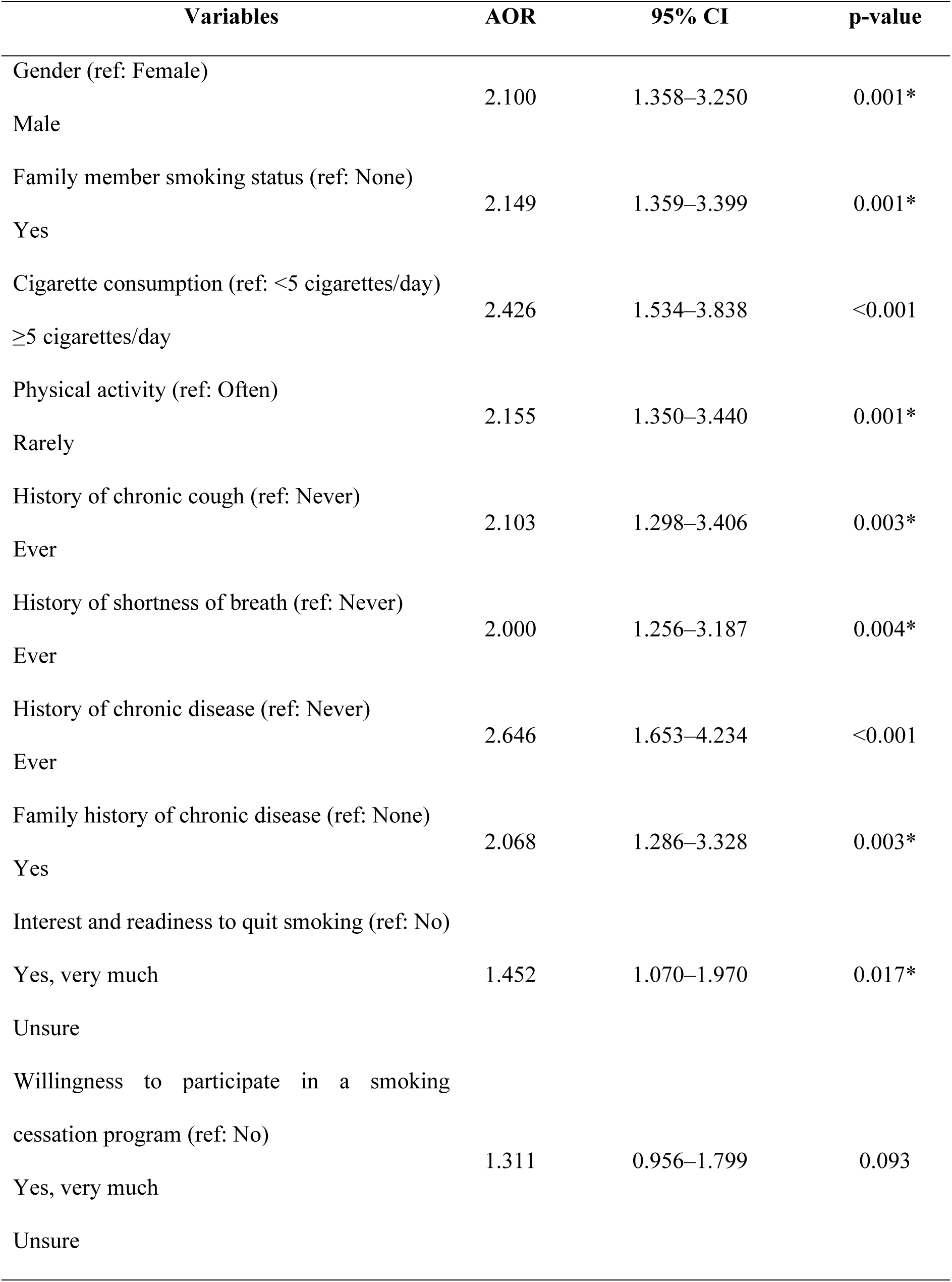
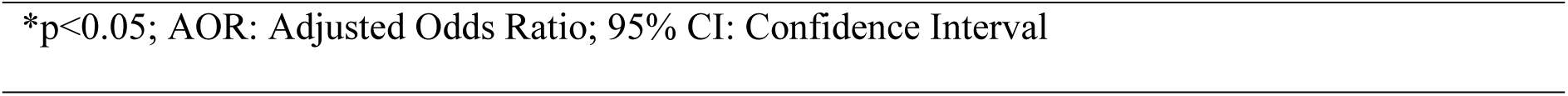
Logistic regression analysis for early detection of chronic disease risk and smoking cessation efforts (n=563).

| Variables | AOR | 95% CI | p-value |
| --- | --- | --- | --- |
| Gender (ref: Female) |  |  |  |
| Male | 2.100 | 1.358–3.250 | 0.001* |
| Family member smoking status (ref: None) |  |  |  |
| Yes | 2.149 | 1.359–3.399 | 0.001* |
| Cigarette consumption (ref: <5 cigarettes/day) |  |  |  |
| ≥5 cigarettes/day | 2.426 | 1.534–3.838 | <0.001 |
| Physical activity (ref: Often) |  |  |  |
| Rarely | 2.155 | 1.350–3.440 | 0.001* |
| History of chronic cough (ref: Never) |  |  |  |
| Ever | 2.103 | 1.298–3.406 | 0.003* |
| History of shortness of breath (ref: Never) |  |  |  |
| Ever | 2.000 | 1.256–3.187 | 0.004* |
| History of chronic disease (ref: Never) |  |  |  |
| Ever | 2.646 | 1.653–4.234 | <0.001 |
| Family history of chronic disease (ref: None) |  |  |  |
| Yes | 2.068 | 1.286–3.328 | 0.003* |
| Interest and readiness to quit smoking (ref: No) |  |  |  |
| Yes, very much | 1.452 | 1.070–1.970 | 0.017* |
| Unsure |  |  |  |
| Willingness to participate in a smoking<br>cessation program (ref: No) |  |  |  |
| Yes, very much | 1.311 | 0.956–1.799 | 0.093 |
| Unsure |  |  |  |

\*p<0.05; AOR: Adjusted Odds Ratio; 95% CI: Confidence Interval

Students who smoke ≥5 cigarettes per day are 2.43 times more likely to have high cotinine levels (AOR=2.426; 95% CI: 1.534–3.838; p<0.001) compared to those who smoke <5 cigarettes per day. This reinforces the evidence that the higher the intensity of smoking, the higher the cotinine levels in the body, reflecting a higher level of nicotine exposure. Students who rarely exercised were 2.16 times more likely to have high cotinine levels (AOR=2.155; 95% CI: 1.350–3.440; p=0.001) compared to students who exercised frequently. This condition indicates that a less active lifestyle may be associated with other unhealthy behaviors, including smoking (Table 3).

Students who have experienced chronic coughing are 2.10 times more likely to have high cotinine levels (AOR=2.103; 95% CI: 1.298–3.406; p=0.003) than those who have never experienced it. This indicates a real physiological impact of nicotine exposure on the respiratory tract, even in adolescents. Students with a history of shortness of breath are also 2 times more likely (AOR=2.000; 95% CI: 1.256–3.187; p=0.004) to have high cotinine levels compared to students without a history of shortness of breath. This reinforces the link between cigarette exposure and impaired lung function at a young age (Table 3).

Students with a history of chronic diseases (such as asthma, bronchitis, or mild hypertension) have a 2.65 times greater risk of having high cotinine levels (AOR=2.646; 95% CI: 1.653–4.234; p<0.001). This suggests that nicotine exposure may exacerbate chronic health conditions or be a risk factor for early comorbidity. Students with a family history of chronic diseases are also 2.07 times more likely (AOR=2.068; 95% CI: 1.286–3.328; p=0.003) to have high cotinine levels than those without such a history. These findings indicate that environmental and family genetic factors contribute to cigarette exposure and the risk of chronic diseases (Table 3).

Students who expressed doubts about quitting smoking were 1.45 times more likely to have high cotinine levels (AOR=1.452; 95% CI: 1.070–1.970; p=0.017) compared to students who were completely uninterested in quitting. This indicates that hesitation and ambivalence in motivation to quit smoking are still associated with high nicotine exposure, suggesting the need for a more intensive motivational approach. The factor of willingness to participate in a smoking cessation program did not show a significant relationship (AOR=1.311; 95% CI: 0.956–1.799; p=0.093). Although there was a tendency for an increased likelihood of high cotinine levels among students who expressed doubt, this relationship was not statistically significant, possibly due to differences in perception or limitations in actual participation in the program (Table 3).

The study findings confirm that nicotine exposure among students is influenced by a combination of behavioral factors, family environment, and health conditions. School and family-based intervention programs that emphasize health education, promotion of physical activity, and motivation-based smoking cessation counseling and biomarkers are highly recommended to prevent the risk of chronic diseases in the future.

## Discussion

The results of the study show that cotinine levels, as a biomarker of nicotine exposure, were found to be high in most students in Praya Barat District. A total of 67% of respondents had cotinine levels above the normal threshold, indicating significant exposure to nicotine either through active smoking or passive exposure to cigarette smoke in the environment. The study findings reinforce the evidence that adolescents are a group vulnerable to tobacco exposure, both due to their own behavior and social and family influences (Marques *et al*., 2021).

Multivariate analysis results show that male students have a 2.1 times higher risk of having high cotinine levels than females (p=0.001). The results of this study are in line with a study by McGinnis et al., which reported that males have a higher prevalence of smoking behavior and nicotine exposure than females due to social, cultural, and gender norms that are more permissive of smoking behavior among males (McGinnis *et al*., 2022). A study conducted by Ekawati et al. states that in Indonesia, the prevalence of smoking among adolescent males reaches more than 30%, which is much higher than that among females (Ekawati *et al*., 2024).

Students who have family members who smoke are 2.15 times more likely to have high cotinine levels than students without family members who smoke (p=0.001). The findings of this study are consistent with those of Fernandes et al., which confirm that exposure to secondhand smoke in the home environment is the main source of increased cotinine levels in children and adolescents (Fernandes *et al*., 2020). In addition, role models and family norms also play a role in shaping adolescent smoking behavior. Families who actively smoke create an environment that is permissive towards tobacco consumption, thereby increasing the risk of children being exposed to or trying smoking (Sim and Park, 2021).

Studies show that students who smoke ≥5 cigarettes per day are 2.43 times more likely to have high cotinine levels (p<0.001). The positive relationship between smoking intensity and cotinine levels has been proven in other studies. Dai et al. mention that cotinine levels increase proportionally with the frequency and number of cigarettes consumed, because nicotine metabolism is converted to cotinine in the body at a relatively stable ratio (Dai *et al*., 2024). Thus, cotinine levels can be considered a quantitative biological marker that reflects the degree of actual tobacco exposure.

Students who rarely engage in sports activities have a 2.16 times greater risk of having high cotinine levels (p=0.001). The results of this study are in line with research by Zhang et al., which found that adolescents with sedentary lifestyles are more likely to smoke and consume other addictive substances (Zhang *et al*., 2022). Low physical activity is often associated with stress, anxiety, and unhealthy social interactions, which are factors that encourage smoking behavior. Conversely, regular exercise has been shown to increase nicotine metabolism and accelerate the removal of toxic substances from the body, as well as increase resistance to nicotine addiction through dopamine regulation (Öksüz *et al*., 2022).

The analysis also showed a significant relationship between high cotinine levels and a history of chronic cough (p=0.003) and shortness of breath (p=0.004). These results indicate that nicotine exposure has a direct impact on the respiratory system of adolescents. A study conducted by Dai et al. states that increased cotinine levels are closely related to decreased lung function, increased incidence of chronic bronchitis, and upper respiratory tract disorders in adolescents (Dai *et al*., 2022). Exposure to cigarette smoke can irritate the bronchial mucosa, increase mucus secretion, and decrease oxygen diffusion capacity in the lungs, even with passive exposure.

Students with a history of chronic disease had a 2.65 times higher risk of high cotinine levels (p<0.001), while those with a family history of chronic disease also showed a 2.07 times higher risk (p=0.003). The study findings support the literature by Benowitz et al., which shows that exposure to nicotine and cigarette smoke are important risk factors for the development of chronic diseases such as hypertension, coronary heart disease, and COPD (Benowitz *et al*., 2023). In addition, a family history of disease may indicate cross-generational exposure to tobacco, either through smoking habits or genetic predisposition to nicotine metabolism.

Students who stated that they were “unsure about quitting smoking” were 1.45 times more likely to have high cotinine levels (p=0.017). These results indicate that motivation to quit smoking is related to the level of nicotine exposure. A study by Gale et al. explains that individuals with low or ambivalent motivation to change their behavior tend to maintain their smoking habits even though they are aware of the negative effects (Gale *et al*., 2022). The low readiness to quit smoking among adolescents is often caused by peer influence, lack of family support, and a lack of effective educational programs.

Although statistically insignificant (p=0.093), the trend shows that students who expressed willingness to participate in a smoking cessation program had relatively high cotinine levels (65%). This may indicate that they are aware of their level of dependence and have the potential motivation to change. A study by Rensch et al. shows that smoking cessation interventions involving a motivational approach and monitoring of biomarkers (such as cotinine levels) can increase the effectiveness of smoking cessation programs in adolescents (Rensch *et al*., 2023).

Overall, the results of this study illustrate that nicotine exposure among students is a serious public health issue, influenced by behavioral, environmental, and physiological factors. The use of cotinine biomarkers has been proven effective in objectively identifying tobacco exposure, even in individuals who do not identify as active smokers. The study’s findings reinforce the Centers for Disease Control and Prevention (CDC) recommendations encouraging the use of biomarker detection in tobacco exposure surveillance among young populations (Anic *et al*., 2022).

Recommended interventions include strengthening smoke-free school programs, educating about the dangers of tobacco, increasing the role of families in controlling smoke-free environments, and developing biomarker-based smoking cessation programs and motivational counseling. This integrated approach has the potential to reduce nicotine exposure from an early age and reduce the risk of chronic diseases in the future.

### Conclusion

The study findings confirm that high cotinine levels in students are influenced by various factors, particularly male gender, having family members who smoke, high cigarette consumption, lack of physical activity, and a history of respiratory disorders and chronic diseases. These results are consistent with various global literature confirming that nicotine exposure in adolescence plays an important role in increasing the risk of chronic diseases later in life. Therefore, biomarker-based approaches such as cotinine need to be integrated into early detection policies and smoking prevention programs among adolescents.

## Artificial intelligence utilization for article writing

Using Grammarly for Punctuation and Grammar Correction in manuscript.

## Authors’ contributions

MH was responsible for conceptualization, writing, design, analysis, interpretation of results, manuscript submission, interpretation of results, and was a major contributor to the manuscript. BBBS and PP were responsible for study authorization, Project administration, data collection, and correcting the study results. MH, BBBS, and PP revised the final manuscript. All authors take full responsibility for the contents of this paper. All the authors have read and approved the final version of the manuscript.

## Availability of data and material

The datasets used in the current study are available upon reasonable request.

## Data Availability

The data underlying the results presented in the study are available from

## Acknowledgements

The authors would like to express their deepest gratitude to the DPPM, Ditjen Risbang, Kemendiktisaintek, Indonesia, for funding this study. In addition, the authors would like to thank the subjects who were willing to participate and colleagues who provided advice and input related to this study.

